# Evolving priorities in malaria research in Indonesia (2015–2025): A scoping review and bibliometric analysis

**DOI:** 10.64898/2026.08.13.26360377

**Authors:** Budi Setiawan, Jonathan M. Cooper, Julien Reboud

## Abstract

**Background:** Malaria remains a major public health challenge in Indonesia, with transmission concentrated in several eastern regions. A systematic overview of national research activity is needed to show how scientific priorities have changed and whether the evidence base is aligned with elimination needs.

**Methodology/Principal findings:** We conducted a scoping review and bibliometric analysis of Scopus-indexed malaria publications from 2015 to 2025 following PRISMA-ScR guidance. Records were screened for an Indonesian institutional affiliation or an explicit Indonesia-specific study component. Publication trends, co-authorship, keyword networks, citation patterns, thematic evolution, and exploratory forecasts were analysed. Keyword text was represented using term frequency–inverse document frequency and grouped with K-means clustering. The initial candidate set contained 592 publications; four false-positive records were excluded, leaving 588 eligible Indonesia-related publications. Of these, 583 had an Indonesian affiliation identifiable in the exported Scopus metadata and 586 had usable keyword metadata for thematic analysis. Ten clusters were identified. Environmental and Community-Based Studies was the largest theme (110/586, 18.77%), followed by Plasmodium Species and Clinical Parasitology (85/586, 14.51%), Molecular Diagnostics (76/586, 12.97%), Treatment and Antimalarial Drugs (69/586, 11.77%), and Health Systems and Malaria Control Programs (67/586, 11.43%). Treatment-related research had the greatest total and mean citation impact. Publication output accumulated continuously without a clear plateau. Forecasts suggested the strongest continued growth in environmental and community-based research, while treatment and health-system themes were comparatively stable.

**Conclusions/Significance:** Indonesian malaria research became more diverse while retaining strong clinical and treatment foundations. Future research planning should balance biomedical priorities with environmental, community, diagnostic, and implementation research. Forecasts are exploratory and should not be interpreted as precise predictions.

**Author summary:** Malaria remains a major health problem in parts of Indonesia, especially in eastern provinces. We examined how malaria research related to Indonesia changed between 2015 and 2025. We reviewed 588 eligible publications indexed in Scopus and used publication records, author links, keywords, citations, and annual trends to identify the main areas of research. We found that research output increased overall but varied considerably from year to year. Clinical studies of malaria parasites and antimalarial treatment remained important, while environmental and community-based research became the largest area. Molecular diagnostic and genetic studies also attracted substantial scientific attention. Research using artificial intelligence was present but still formed only a small part of the overall evidence base. Our exploratory forecasts suggest that environmental and community research may continue to grow most strongly, whereas treatment and health-system research may remain relatively stable. These findings show that malaria research related to Indonesia has become more diverse rather than reaching a stable or mature endpoint. We believe that future planning should maintain strong clinical and treatment research while giving greater attention to environmental conditions, community participation, diagnostic implementation, and health-system needs that directly support malaria elimination.

## Introduction

Malaria remains a major global health challenge, with a burden that has shown limited reduction in recent years despite substantial control efforts. In 2014, an estimated 198 million malaria cases and 584,000 deaths were recorded worldwide [1]. Global case numbers have continued to rise, reaching approximately 229 million in 2019 [2] and increasing to 241 million in 2020 due to widespread disruptions in malaria services linked to the COVID-19 pandemic [3]. Subsequent World Malaria Reports estimated 247 million cases in 2021 [4], 249 million in 2022 [5], and 263 million cases with roughly 597,000 deaths in

2023 [6]. In 2024, the estimated number of malaria cases increased further to 282 million [7]. Although mortality declined to its lowest level since before the pandemic, the upward trend in cases signals ongoing challenges in sustaining global progress.

The highest burden continues to fall on the WHO African Region, which accounted for approximately 88% of global cases in 2024 [7]. In the Western Pacific Region, 2.4 million malaria cases were reported across nine endemic countries (1% of cases globally), with Papua New Guinea contributing the largest number, followed by Indonesia and the Solomon Islands [7]. Indonesia is therefore among the countries in which persistent transmission remains concentrated in specific high-burden regions.

The gradual rise in global cases is concerning, especially following decades of substantial reductions achieved through vector control, improved diagnostics, and preventive strategies. Reversals in malaria trends emphasize the need for proactive, detailed assessments of the drivers contributing to recent increases[8,9]. In Indonesia, structural and socioeconomic factors present considerable challenges. In 2023, 9.4% of the population lived below the poverty line, 7.3% in urban areas and 12.2% in rural areas[10], while approximately 65% of the population resides in malaria-endemic provinces such as Papua, West Papua, and East Nusa Tenggara [8]. These areas also have Human Development Index (HDI) values below the national average, underscoring the relationship between poverty, limited health access, and sustained malaria transmission[11].

Malaria has been documented in Indonesia since at least 1900, with the earliest reports by Robert Koch in Central Java and Jakarta [12]. Control efforts expanded during the Global Malaria Eradication Programme, but political instability such as the 1965 conflict hampered progress [13]. Today, limited access to primary healthcare remains a major barrier in endemic provinces. Between 14% and 54% of Public Health Centers in Central Papua, West Papua, and Southwest Papua operate without a physician [14]. Nationally, Indonesia has only 5.7 medical laboratory technicians (MLTs) per 100,000 population[14], far below neighboring countries such as Malaysia (23.9/100,000) [15] and Singapore (42.1/100,000) [16]. This shortage contributes to delays in diagnosis, reduced surveillance sensitivity, and high workloads for existing laboratory personnel.

Despite sustained control efforts, malaria remains entrenched in several regions. In Papua, the contribution to national malaria cases has increased dramatically from 40% to 90% between 2010 and 2019 [17]. In 2021, Indonesia reported 94,610 cases across 167 of its 514 districts [18], rising sharply to 415,140 cases in 2022 [19]. The eastern provinces including Papua, West Papua, Maluku, North Maluku, and East Nusa Tenggara consistently account for 75–80% of national cases [20]. By 2023, only 8 of the country’s 38 provinces were predicted to have achieved malaria elimination status. The Annual Parasite Index (API) fluctuated between 0.8 and 1.6 per 1,000 population from 2019 to 2022 [14], reflecting persistent transmission.

At the same time, significant advancements in malaria diagnostics, surveillance, and vector control have transformed the scientific landscape. Novel biomarkers, next-generation sequencing, proteomics, and metabolomics have improved diagnostic speed and accuracy[21]. Rapid diagnostic tests (RDTs) and molecular assays enable earlier treatment and reduced transmission. Vector control strategies traditionally built on insecticide-treated nets (ITNs) and indoor residual spraying (IRS) are increasingly supplemented by next-generation insecticides and genetic innovations such as gene drive systems and symbiont-based control approaches [22,23]. The RTS,S/AS01 vaccine, although modest in efficacy, represents a major milestone in malaria prevention and has been introduced into routine childhood immunization in some countries [24]. Nevertheless, emerging drug and insecticide resistance, climate-driven changes in vector ecology, and limited numbers of trained entomologists continue to hinder progress [25]. New tools, including connected diagnostics and digital surveillance systems, offer opportunities to strengthen real-time monitoring, particularly in resource-constrained settings [21].

Although numerous studies have examined malaria epidemiology, diagnostics, treatment, and vector dynamics in Indonesia, a comprehensive synthesis of national research trends over the past decade is still lacking. Understanding research output is essential for identifying strengths, gaps, and evolving priorities that can inform the country’s malaria elimination strategies. This scoping review and bibliometric analysis address this gap by mapping ten years of malaria research in Indonesia. The review provides an integrated assessment of publication trends, thematic evolution, and research impact, while identifying areas requiring further investigation particularly in laboratory diagnostics and precision-based approaches. Ultimately, this study situates Indonesia’s scientific progress within broader global efforts toward malaria elimination and seeks to guide future policy and research directions[26–28].

This review addresses three key questions: first, what the publication trends and dominant research themes are in malaria studies conducted in Indonesia between 2015 to 2025; second, which studies demonstrate the greatest intellectual influence, as reflected by citation patterns; and third, what research gaps and future priorities emerge that may support Indonesia’s malaria elimination agenda.

## Materials and methods

### Data source

Bibliographic records related to malaria were retrieved from the Scopus database in January 2026. Scopus was selected as the sole data source due to its broad coverage of peer-reviewed biomedical and health sciences literature, as well as its comprehensive citation metadata, which is advantageous for bibliometric analysis [29,30]. Scopus is widely regarded as one of the most extensive multidisciplinary databases available [31]. We acknowledge that restricting the search to a single database may introduce selection bias, and future studies could incorporate multiple sources to achieve more exhaustive coverage of malaria research output.

### Study design

#### Protocol and registration

This review was not registered, and no protocol was prepared. To guide the scope and boundaries of this review, a structured framework was applied to ensure clarity in defining what types of evidence would be examined and how the mapping process would be conducted. Because this study adopts a scoping review approach aimed at characterizing the breadth, themes, and evolution of Indonesian affiliated research on malaria, the Population–Concept–Context (PCC) framework recommended by the Joanna Briggs Institute (JBI) was used [32]. The PCC framework offers a suitable structure for reviews that seek to map existing knowledge rather than evaluate intervention effects, and it allows the analytical focus to be aligned with the bibliometric and text-mining methods employed in this study. Based on this rationale, the PCC elements for the present review were defined as follows:

**Population (P):** Scopus-indexed publications related to malaria research in Indonesia and published between 2015 and 2025. Eligible records included publications with an Indonesian institutional affiliation and multicountry or regional studies in which Indonesia was explicitly represented as a study setting, sampling location, population, data source, policy or intervention context, or analytical unit.

**Concept (C):** The application of bibliometric mapping and text-mining approaches including TF-IDF analysis, unsupervised machine learning clustering, and trend prediction to characterize research themes, growth patterns, research themes impact, and emerging directions in malaria research.

**Context (C):** The global scientific landscape as represented Scopus database, with projections extending to 2030 to understand future trajectories in Indonesian contributions to malaria research. The overall review and bibliometric workflow is summarised in Fig 1.

**Fig 1.**
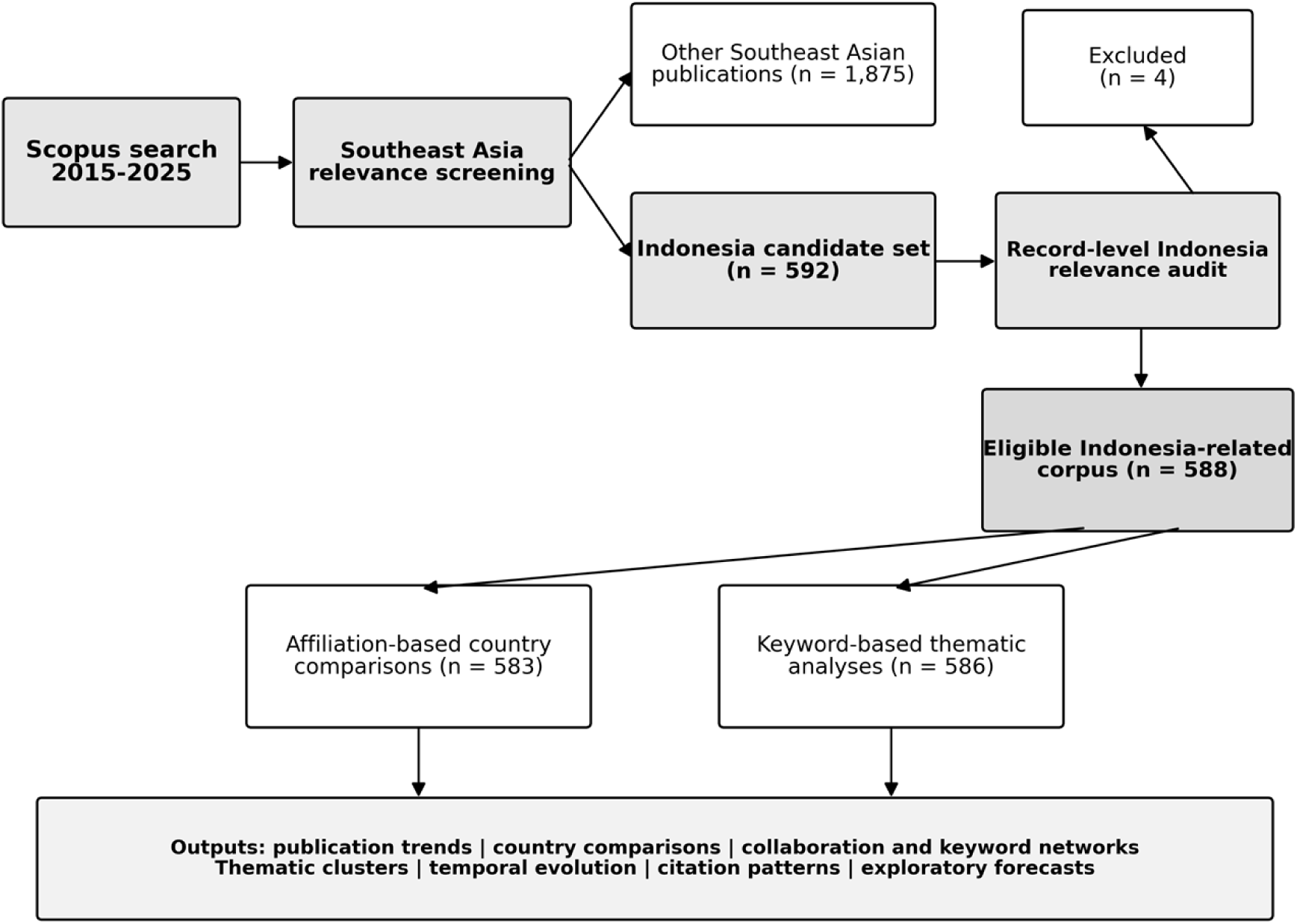
Bibliometric study workflow.

### Eligibility criteria

Publications were eligible if they were published between 1 January 2015 and 31 December 2025, indexed in Scopus, written in English or Indonesian, and substantively related to malaria research in Indonesia. Indonesia-related publications were defined as studies meeting at least one of the following criteria: an Indonesian institutional affiliation; explicit inclusion of Indonesia as a study setting, sampling location, study population, data source, policy or intervention context, or analytical unit; or inclusion of Indonesian data within a multicountry or regional analysis. Duplicate, retracted, and clearly non-Indonesia records were excluded. Records lacking a title or abstract were excluded from the review. Keyword-based thematic analysis additionally required at least one usable author keyword or index keyword. The search was executed on 7 January 2026.

### Study selection and data extraction

Bibliographic metadata were exported from Scopus in comma-separated values (CSV) format. Extracted variables included title, abstract, author keywords, index keywords, author names and affiliations, publication year, source title, citation counts, and unique document identifiers. A final record-level audit was conducted to verify substantive relevance to Indonesia. Four records without an Indonesian affiliation or Indonesia-specific study component were removed from the initial 592-record candidate set, yielding 588 eligible Indonesia-related publications. Of these, 583 records had an Indonesian affiliation identifiable in the exported Scopus Affiliations field and were used for reproducible affiliation-based country comparisons. Two eligible records lacked both author keywords and index keywords; these records were retained in descriptive analyses but excluded from keyword-based modelling, resulting in a thematic corpus of 586 publications. Affiliations were standardised to the parent institution level for institution-based analyses. The variables sought from each record were predefined according to the objectives of the bibliometric and thematic analyses. Title and abstract were retained as bibliographic text describing the publication content. Author Keywords and Index Keywords were treated as author- and database-assigned descriptors of research content, respectively. Author names were used for authorship and co-authorship analyses, while institutional affiliations were used to determine country and institutional contributions. Publication year was defined as the year reported in the Scopus record and was used for annual and temporal analyses. Source title represented the journal in which the publication appeared. Citation count was defined as the number of citations recorded in Scopus at the time of data extraction and was used as a descriptive indicator of scholarly influence. Unique Scopus document identifiers were used to distinguish records and support duplicate checking.

Data charting was performed directly from the structured bibliographic fields exported from Scopus rather than using a separate manually calibrated extraction form. The predefined variables used for charting were title, abstract, author and index keywords, author names, institutional affiliations, publication year, source title, citation counts, and unique document identifiers. Because these variables were obtained from standardized database fields and subsequently processed using reproducible computational workflows, duplicate independent data extraction was not undertaken. Records with ambiguous eligibility, affiliation, or Indonesia-related status were subjected to record-level manual verification before inclusion in the final analytical corpus. No study investigators or corresponding authors were contacted to obtain or confirm additional data, as all analyses were based exclusively on the bibliographic metadata available from Scopus.

Several operational assumptions and simplifications were applied. Country assignment for affiliation-based comparisons relied on explicitly stated country information in the exported Scopus Affiliations field; author nationality was not inferred from names. For the broader Indonesia-related corpus, a publication was considered relevant when it contained either an Indonesian institutional affiliation or an explicit Indonesia-specific study setting, population, sampling location, data source, policy or intervention context, or analytical component. Institutional sub-units were consolidated to their parent institutions when the parent institution was explicitly identifiable. Author Keywords and Index Keywords were combined into a single keyword field for thematic modelling, and records lacking both keyword fields were excluded only from keyword-based analyses. Citation counts were analysed as recorded in Scopus and were not interpreted as direct measures of research quality. No missing keyword values were imputed.

### Search strategy

The search strategy was developed using the Scopus database and executed in January 2026. The bibliometric query employed Boolean operators and field restrictions as follows: Search string:**`( TITLE ( malaria ) OR TITLE ( Plasmodium ) ) AND PUBYEAR > 2014 AND PUBYEAR < 2026 AND ( LIMIT-TO ( DOCTYPE , "ar" )`** ) AND ( LIMIT-TO ( LANGUAGE , "English" ) OR LIMIT-TO ( LANGUAGE , **`"Indonesian" ) )`**

The search covered publications dated from 1 January 2015 to 31 December 2025 and was restricted to peer-reviewed journal articles written in English or Indonesian. Initial records were screened for Southeast Asian relevance. The Indonesia-related corpus included publications with an Indonesian institutional affiliation and multicountry or regional studies in which Indonesia was explicitly represented as a study setting, sampling location, population, data source, policy or intervention context, or analytical unit. For regional country comparisons, a stricter and reproducible metadata-based rule was applied: a publication was counted for Indonesia only when the exported Scopus Affiliations field contained at least one Indonesian institutional affiliation. This distinction allowed thematic analyses to capture research directly relevant to Indonesia while maintaining a consistent affiliation-based definition for national publication comparisons.

### Data analysis

Scopus records were cleaned and curated before bibliometric processing. Descriptive analyses used the 588-publication eligible Indonesia-related corpus, whereas affiliation-based country comparisons used the 583 records with an Indonesian affiliation identifiable in the exported Scopus Affiliations field. Keyword-based analyses used 586 eligible publications with usable author or index keyword metadata. Charted bibliographic data were summarized descriptively according to the analytical objective. Publication output was summarized using annual document counts and year-to-year percentage changes, while country, institutional, and thematic distributions were expressed as absolute frequencies and percentages. Citation impact was summarized at the thematic-cluster level using total citations and mean citations per publication. Temporal thematic patterns were calculated as the annual number and proportion of publications assigned to each thematic cluster. Co-authorship and keyword relationships were summarized through network-based measures and visualized using VOSviewer. Keyword information was further reduced to TF–IDF representations for unsupervised clustering, and cluster-level outputs were summarized using document counts, proportions, and dominant terms. No quantitative pooling of clinical outcomes or meta-analysis was performed.

Author Keywords and Index Keywords were combined and transformed using TF–IDF vectorisation with a maximum of 400 features, unigram and bigram terms, a minimum document frequency of 2, a maximum document frequency of 0.8, and English stop words. K-means clustering used k-means++ initialisation, 15 initialisations, a maximum of 300 iterations, and a random seed of 42. Candidate solutions from k = 2 to k = 10 were compared using inertia, silhouette score, Davies–Bouldin index, and Calinski–Harabasz score. VOSviewer was used for complementary co-authorship and keyword co-occurrence visualisation, with minimum thresholds of two publications per author and five occurrences per keyword. Exploratory forecasts for five leading themes were generated by fitting a damped Holt linear-trend model separately to annual publication counts from 2015 to 2025 and projecting values for 2026–2030.

To further examine the structural characteristics and developmental status of research themes, a thematic mapping approach was applied using a quadrant-based visualization. In this analysis, *centrality* represents the relative importance of a theme within the overall research network and was operationalized using co-word importance derived from TF–IDF weighted keyword co-occurrence and network centrality measures. *Density* reflects the internal cohesion and maturity of each theme and was calculated based on the strength of interconnections among keywords within the same cluster. By plotting themes according to their centrality and density values, the thematic map enables classification of research topics into core, emerging, niche, and declining themes, thereby providing insight into the intellectual structure and evolutionary dynamics of malaria research in Indonesia.

### Statistical software and reproducibility

All analyses were conducted in Python version 3.9.13 using pandas 1.5.3, NumPy 1.24.3, scikit-learn 1.3.0, matplotlib 3.7.1, seaborn 0.12.2, SciPy 1.10.1, statsmodels 0.14.0, and NLTK 3.8.1. The analysis code, environment configuration, output tables and figures, and a detailed README are intended to be archived in a public repository to support reproducibility.

### Risk of bias and certainty assessment

Because this review did not evaluate clinical, epidemiological, or health outcomes, no risk-of-bias assessment (e.g., Cochrane RoB, ROBINS-I) or certainty grading (e.g., GRADE) was performed. Reporting bias was not assessed, and meta-analytic measures such as effect estimates, heterogeneity, or sensitivity analyses were not applicable [33].

### Ethical considerations

This study analysed publicly available bibliographic metadata and did not involve human subjects or patient data. No ethics approval was required. All data were used in accordance with Scopus Terms of Use and academic fair use provisions.

## Results

### Study selection and corpus definition

A final record-level relevance audit of the 592 Indonesia-candidate publications identified four false-positive records that had neither an Indonesian institutional affiliation nor a substantive Indonesia-specific study setting, population, data source, policy context, intervention context, or analytical component. Their exclusion yielded a final eligible Indonesia-related corpus of 588 publications.

For affiliation-based country comparisons, 583 publications with at least one Indonesian institutional affiliation identifiable in the exported Scopus Affiliations field were counted as Indonesian research output. The same metadata-based country-assignment rule was applied consistently across Southeast Asian countries.

Keyword-based thematic analyses required usable author or index keyword metadata. Two of the 588 eligible publications lacked both keyword fields and were retained only in descriptive analyses. Consequently, 586 publications were included in TF–IDF vectorisation, thematic clustering, temporal theme analysis, citation analysis by cluster, and forecasting. The different denominators therefore reflect final relevance screening and analysis-specific metadata requirements rather than inconsistencies in the underlying dataset.

Fig 2 presents the PRISMA-ScR flow of record identification, screening, eligibility assessment, and analysis-specific inclusion. Regional screening retained 2,467 Southeast Asian publications, of which 592 were initially classified as Indonesia-related and 1,875 were associated with other Southeast Asian countries.

**Fig 2.**
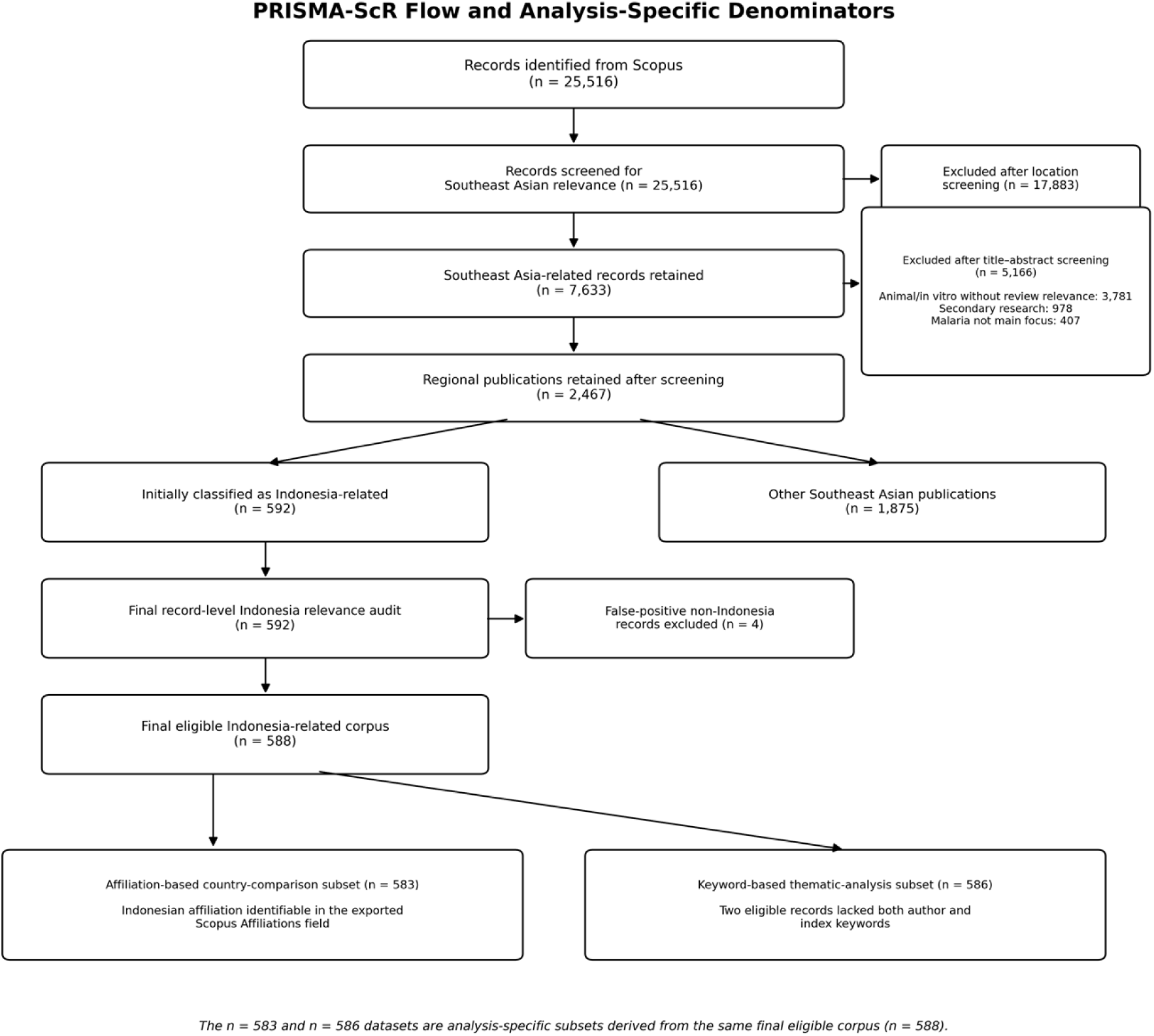
PRISMA-ScR flow diagram for the regional screening process and the analysis-specific Indonesia-related datasets. The final eligible Indonesia-related corpus contained 588 publications; 583 were used for metadata-based affiliation comparisons and 586 for keyword-based thematic analysis.

### Malaria research in Southeast Asia

Fig 3A illustrates the distribution of malaria-related publications across Southeast Asian countries from 2015 to 2025, as well as Indonesia’s contribution in comparison with the overall regional output. Thailand is the most contributor followed by Indonesia and Malaysia. Singapore and Cambodia show moderate levels of publication, while Myanmar, the Philippines, Vietnam, Brunei Darussalam, and Lao PDR contribute relatively few documents over the study period. Panel (b) compares annual malaria publications from Indonesia with the total output from Southeast Asia. Indonesia consistently contributes a proportion of regional malaria research, although its output remains lower than the aggregated Southeast Asia total. The temporal trends of Indonesia and the region appear broadly parallel, with peaks and declines occurring in similar periods. Overall, Fig 3 highlights Indonesia as a key contributor to malaria research in Southeast Asia, while also pointing to opportunities for strengthening research output to support regional malaria elimination efforts.

**Fig 3.**
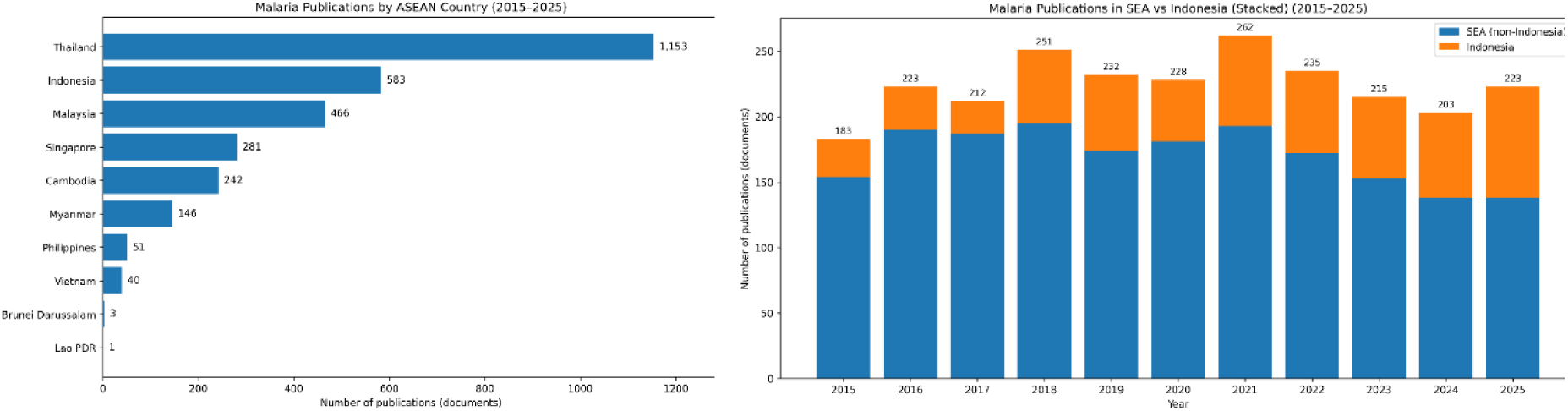
Malaria-related publication output in Southeast Asia and Indonesia, 2015–2025. (A) Cumulative publication output by Southeast Asian country. Thailand had 1,153 publications, Indonesia 583, and Malaysia 466. (B) Annual regional output, with Indonesia shown separately from the combined output of other Southeast Asian countries.

### Co-authorship networks

Fig 4 illustrates the co-authorship networks of malaria research in Southeast Asia (Fig 4A) and Indonesia (Fig 4B) during the period 2015–2025. At the regional level, the author network shows a dense and interconnected structure, with several prominent clusters indicating strong collaborative relationships among researchers across different Southeast Asian countries. A small number of highly connected authors appear to act as central nodes, suggesting their key role in facilitating regional research collaboration and knowledge exchange. In contrast, the Indonesian author network displays a more fragmented structure with fewer cross-cluster connections. Collaboration is largely concentrated within smaller research groups, with limited bridging authors connecting different clusters. This pattern indicates that malaria research collaboration in Indonesia is more locally oriented, with less integration across broader national or regional research networks.

**Fig 4.**
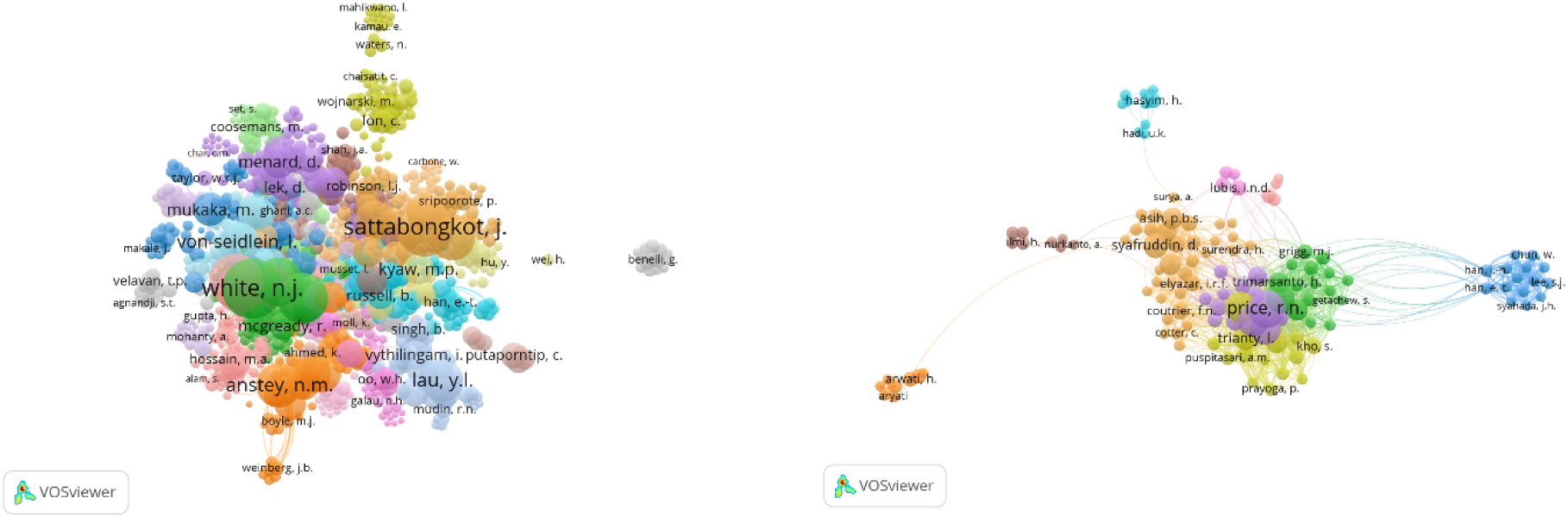
Co-authorship networks in Southeast Asia and Indonesia, 2015–2025. (A) Network among authors contributing to malaria research across Southeast Asia. (B) Network restricted to Indonesia-related publications. Node size indicates publication frequency, lines indicate co-authorship, line thickness indicates collaboration strength, and colours identify collaboration clusters.

### Keyword co-occurrence and thematic analysis

Fig 5 presents the co-occurrence keyword networks of malaria research in Southeast Asia (Fig 5A) and Indonesia (Fig 5B) for the period 2015–2025. At the regional level, the network is characterized by multiple well-defined clusters, indicating a broad thematic landscape encompassing clinical parasitology, vector biology, treatment, and public health–related topics. Frequently occurring keywords form dense hubs, reflecting sustained research attention to core malaria concepts and established research domains across Southeast Asia. The Indonesian keyword network displays a similarly structured core, with high-frequency terms related to malaria, human infection, and Plasmodium species occupying central positions. Several thematic groupings are evident, suggesting active research across clinical, epidemiological, and diagnostic domains. The overall organization of the network highlights the centrality of fundamental malaria-related concepts while also showing thematic diversification within the Indonesian research context.

**Fig 5.**
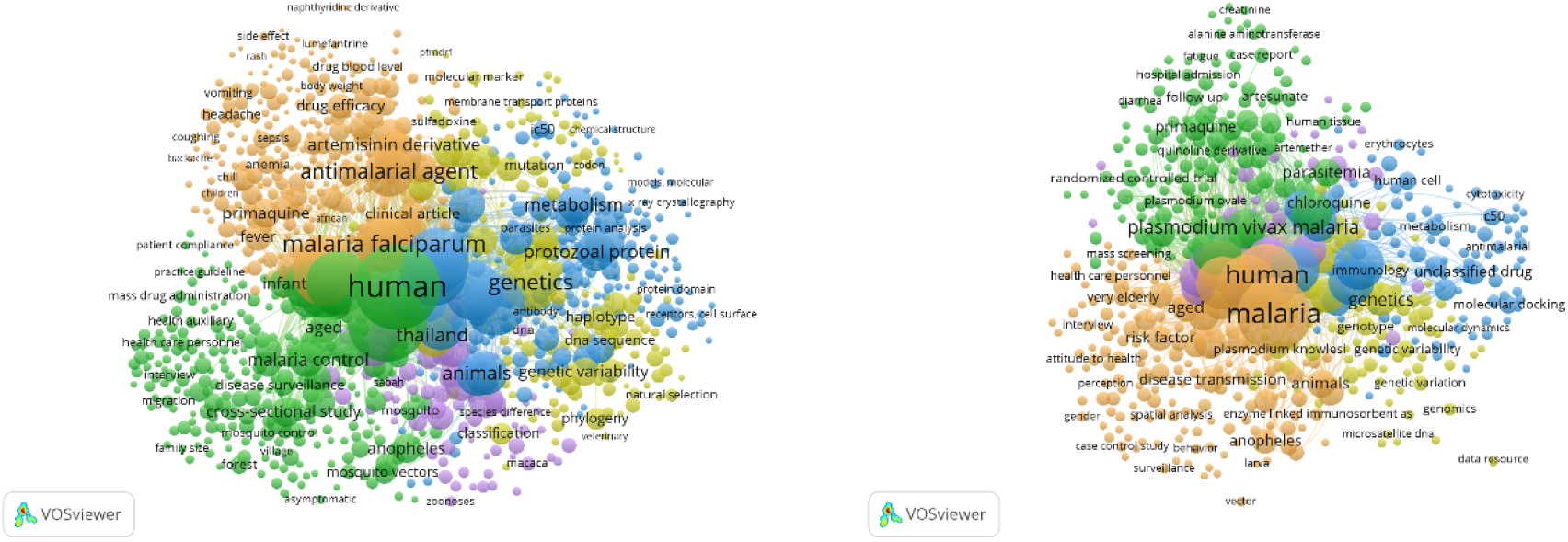
Keyword co-occurrence networks in Southeast Asia and Indonesia, 2015–2025. (A) Network for Southeast Asian malaria publications. (B) Network for Indonesia-related publications. Node size represents keyword frequency, links represent co-occurrence, and colours indicate VOSviewer clusters.

### Clustering analysis

Fig 6 presents the cluster optimisation results for the final 586-publication thematic corpus. The silhouette score reached its highest value at k = 10 (0.059), although the overall values remained low, indicating overlapping thematic boundaries. The Davies–Bouldin index reached its minimum at k = 9 (3.494) and remained relatively low at k = 10 (3.631). Inertia decreased progressively without a distinct elbow, while the Calinski–Harabasz score was highest at k = 2 and declined as k increased. Considering the highest silhouette score, the near-minimum Davies–Bouldin index, and the need to retain sufficient thematic detail, k = 10 was maintained as a pragmatic and interpretable solution rather than an unequivocally optimal solution across all metrics.

**Fig 6.**
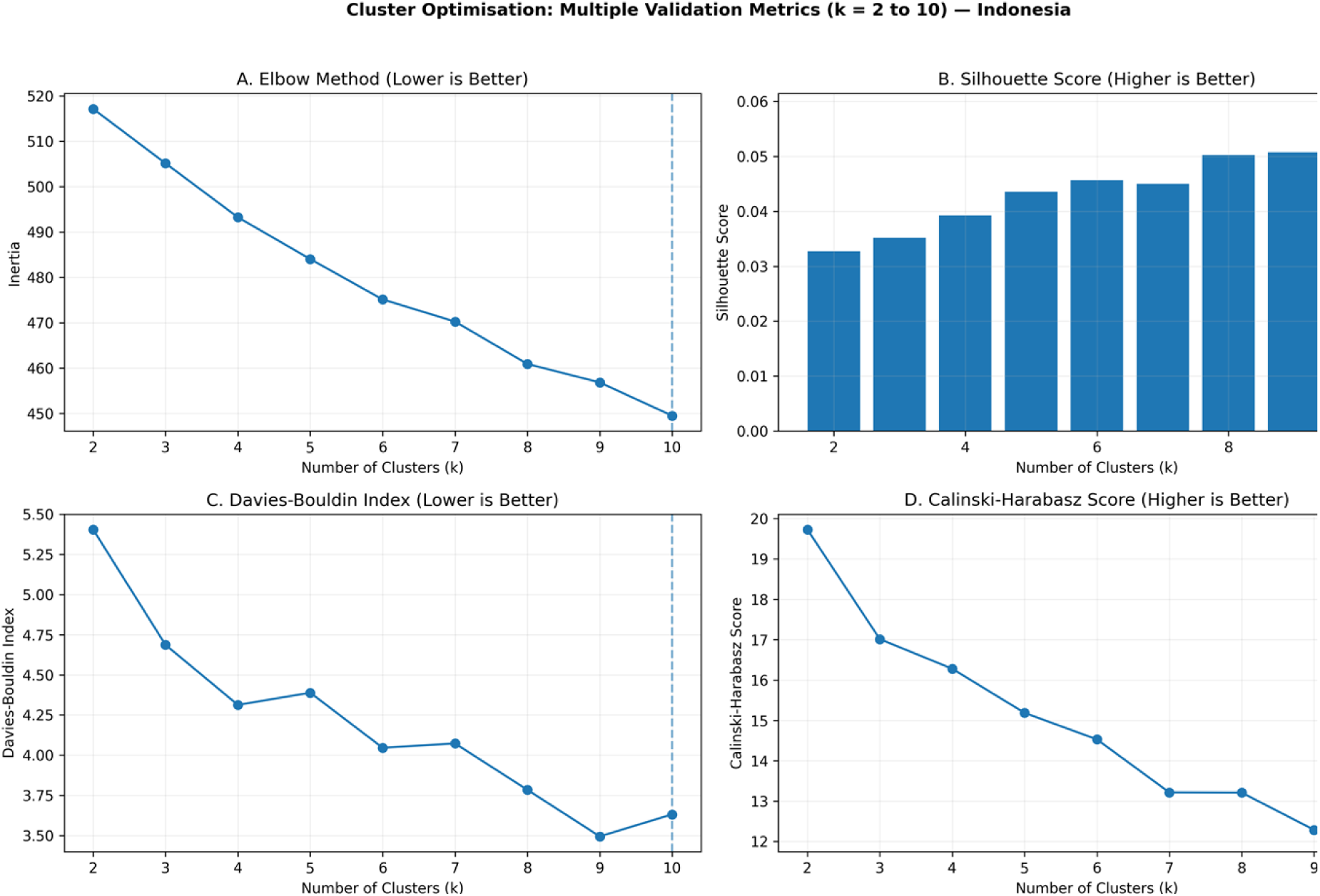
Cluster optimisation for the final 586-publication keyword corpus. The silhouette score was highest at k = 10 (0.059); the Davies–Bouldin index was lowest at k = 9 (3.494) and remained relatively low at k = 10 (3.631). Inertia declined without a distinct elbow, while the Calinski–Harabasz score favoured fewer clusters. The ten-cluster solution was retained as a pragmatic balance between internal validation and thematic resolution.

The final keyword-based corpus comprised 586 eligible Indonesia-related publications distributed across ten thematic clusters (Table 1). Environmental and Community-Based Studies formed the largest cluster (18.77%), followed by Plasmodium Species and Clinical Parasitology (14.51%) and Molecular Diagnostics (PCR-based) (12.97%). Treatment and Antimalarial Drugs accounted for 11.77%, while Health Systems and Malaria Control Programs represented 11.43% of the thematic corpus.

**Table 1.**
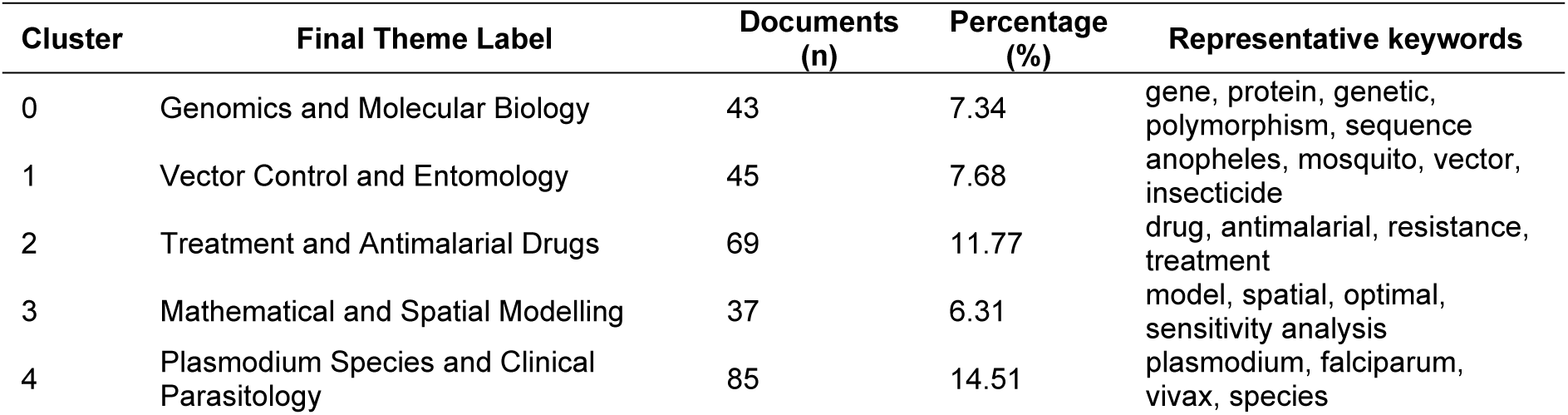
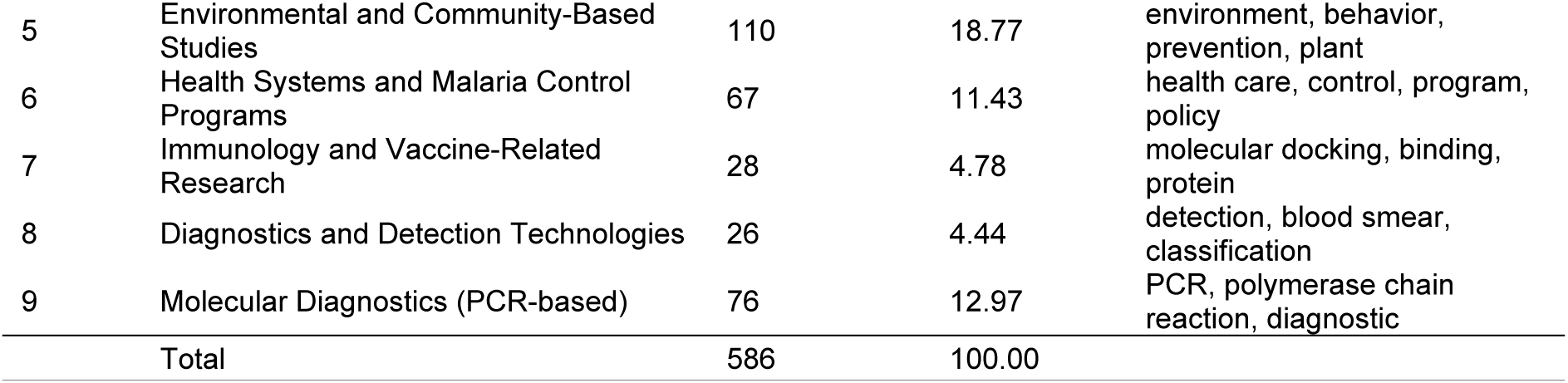
Thematic clusters of malaria research related to Indonesia (2015–2025)

The remaining clusters comprised Vector Control and Entomology (7.68%), Genomics and Molecular Biology (7.34%), Mathematical and Spatial Modelling (6.31%), Immunology and Vaccine-Related Research (4.78%), and Diagnostics and Detection Technologies (4.44%). The ordering of the ten themes was unchanged after the eligibility correction, and the largest change in cluster share was 0.25 percentage points.

Overall, the thematic distribution demonstrates a research landscape that balances applied public health priorities with laboratory-based and methodological studies. While Indonesia shares broad thematic similarities with the wider Southeast Asian research landscape particularly in areas related to parasite biology, treatment, and control strategies the relative emphasis differs across themes.

### Temporal evolution

Fig 7 illustrates annual and cumulative publication dynamics for the final 588-publication eligible Indonesia-related corpus. Annual output increased overall but fluctuated substantially, declining from 33 publications in 2016 to 25 in 2017 (−24.2%), increasing to 55 in 2018 (+120.0%; Δn = 30), rising from 47 in 2020 to 68 in 2021 (+44.7%; Δn = 21), and reaching the highest annual output of 84 publications in 2025 (+31.3%; Δn = 20 compared with 2024).

**Fig 7.**
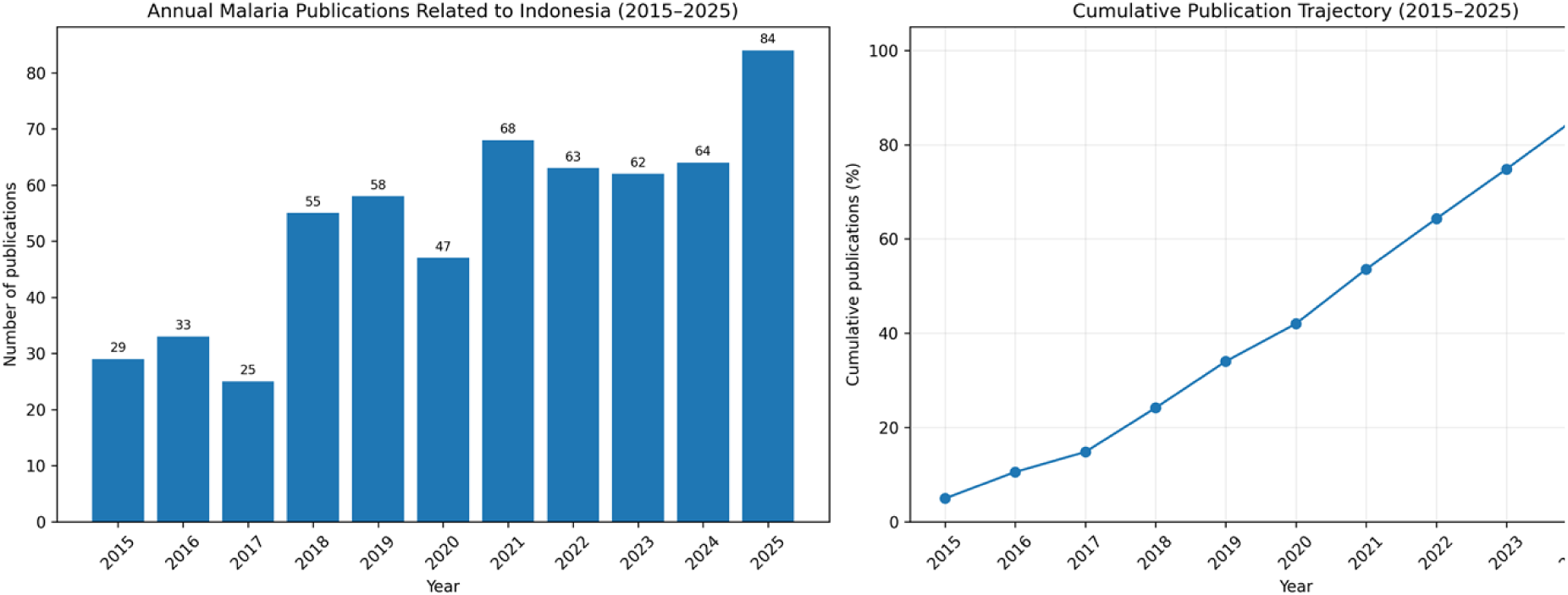
Publication dynamics of malaria research related to Indonesia, 2015–2025. (A) Annual publication output in the 588-publication eligible corpus. (B) Cumulative publication trajectory, which continued to rise through 2025 without a clear plateau.

The cumulative trajectory shows continuous knowledge accumulation rather than a clearly defined sigmoid curve or plateau. Approximately 10.5% of the eligible publications had appeared by 2016, 53.6% by 2021, and 85.7% by 2024. The continued rise through 2025 is therefore more appropriately interpreted as ongoing expansion and thematic diversification than as evidence of a fully mature or stabilised research field.

Fig 8 shows the annual percentage distribution of themes within the final 586-publication thematic corpus. During 2015–2017, Treatment and Antimalarial Drugs accounted for an average of 22.3% of annual themed publications, Plasmodium Species and Clinical Parasitology for 20.6%, and Molecular Diagnostics (PCR-based) for 19.2%.

**Fig 8.**
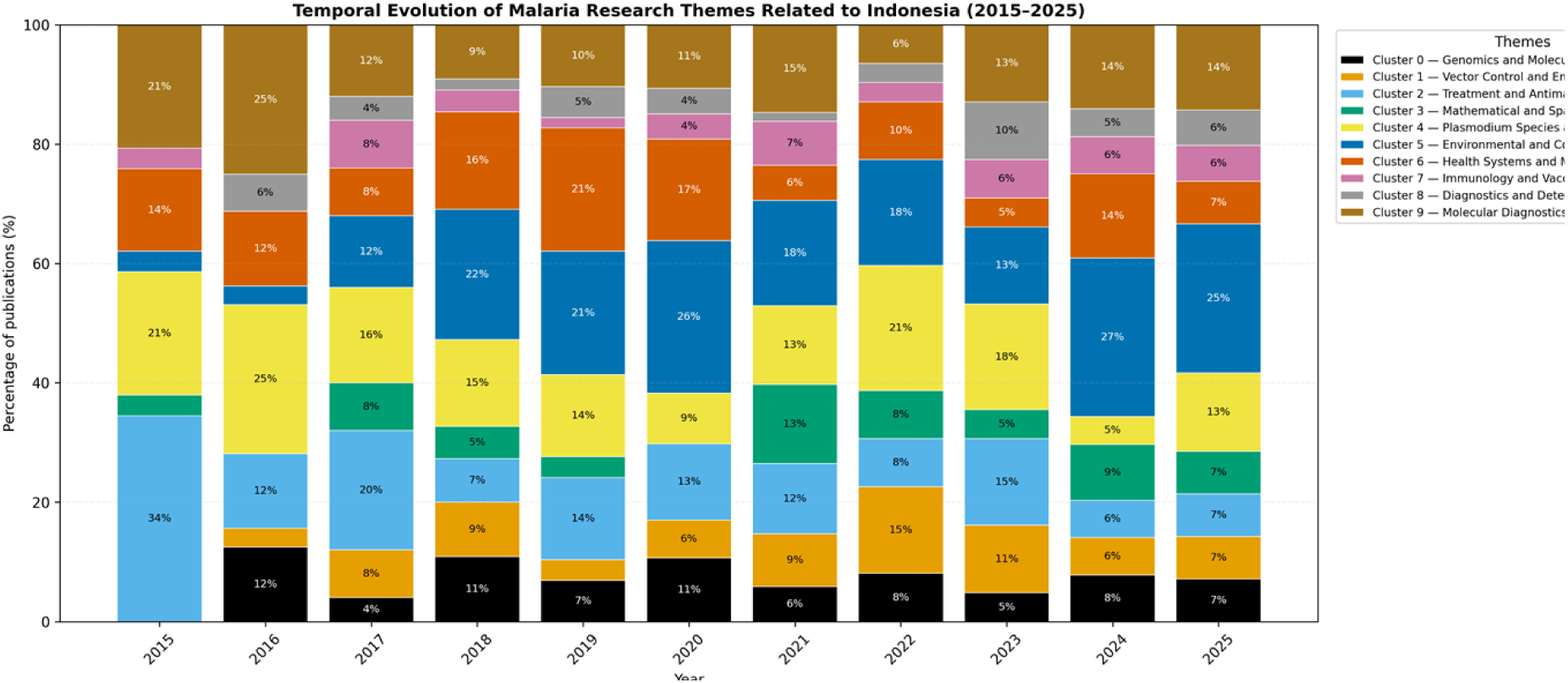
Temporal evolution of malaria research themes related to Indonesia (2015–2025). Each bar presents the annual percentage distribution of the final 586-publication thematic corpus. Treatment and Antimalarial Drugs accounted for 34% in 2015, while Plasmodium Species and Clinical Parasitology and Molecular Diagnostics each accounted for 25% in 2016. Environmental and Community-Based Studies represented approximately 22% in 2018, 21% in 2019, 26% in 2020, 27% in 2024, and 25% in 2025. Percentages show relative annual composition and do not indicate absolute declines in any theme.

Genomics and Molecular Biology represented 5.5%, Environmental and Community-Based Studies 6.2%, and Vector Control and Entomology 3.7%.

During 2018–2021, the thematic distribution became more diverse. Environmental and Community-Based Studies accounted for an average of 21.4% of annual themed publications, followed by Health Systems and Malaria Control Programs at 15.0%.

Plasmodium Species and Clinical Parasitology represented 12.5%, Molecular Diagnostics (PCR-based) 11.2%, and Genomics and Molecular Biology 8.6%. These changes indicate redistribution of thematic emphasis rather than disappearance of biomedical or molecular research.

During 2022–2025, Environmental and Community-Based Studies remained the largest theme, averaging 20.6% of annual themed publications. Plasmodium Species and Clinical Parasitology represented 14.1%, Molecular Diagnostics (PCR-based) 11.9%, Vector Control and Entomology 9.8%, Health Systems and Malaria Control Programs 8.9%, Treatment and Antimalarial Drugs 9.0%, and Genomics and Molecular Biology 7.0%.

Overall, the temporal pattern suggests a rebalancing of malaria research in Indonesia from an early concentration on biomedical, clinical, and molecular themes toward a broader combination of environmental, community-based, health-system, vector-control, and diagnostic research. This pattern is consistent with increasing attention to the multidisciplinary requirements of malaria control and elimination. However, the percentages represent the relative annual distribution of publications and should not be interpreted as evidence of a decline in the absolute number of publications within any particular theme.

Fig 9 and Table 2 together illustrate changes in the structural position of major malaria research themes in Indonesia between 2015–2019 and 2020–2025. During the earlier period, the human and clinical research theme occupied the motor-theme quadrant, indicating relatively high centrality and density and suggesting that it was both well developed and strongly connected to the broader research field. The treatment and antimalarial theme was positioned near the intersection of the thematic quadrants, reflecting an intermediate structural role, while the genomics, animal, and Anopheles-related theme remained relatively peripheral.

**Fig 9.**
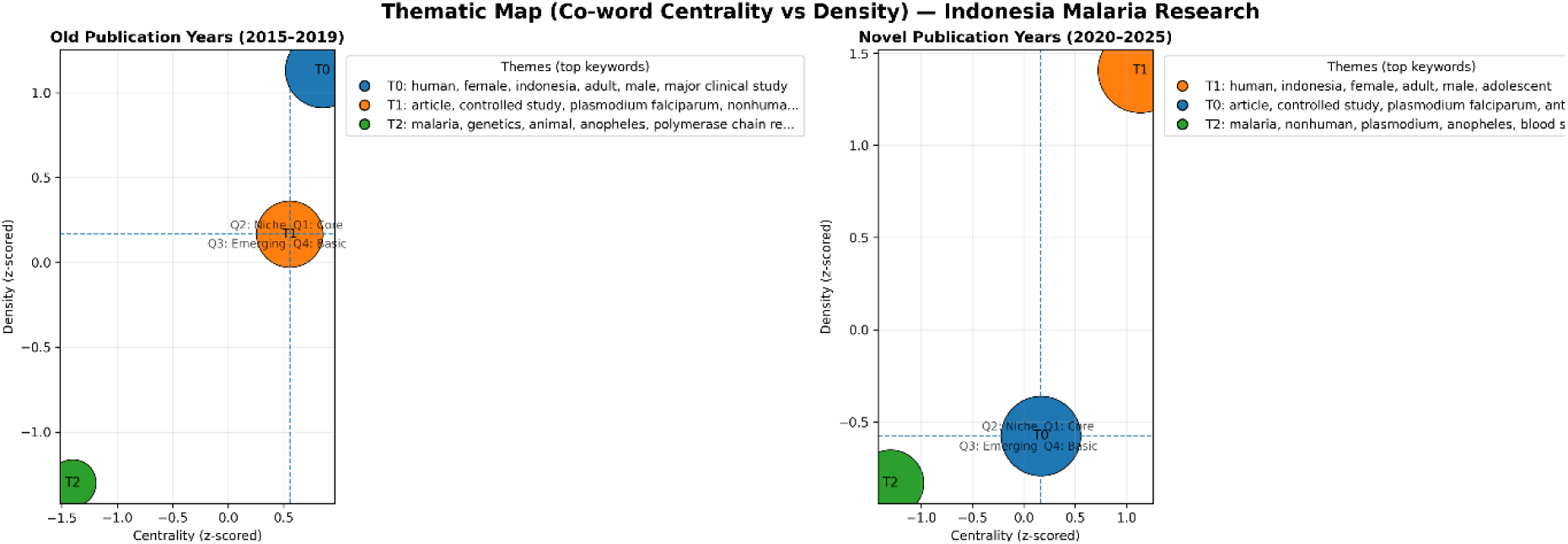
Quadrant plot for malaria research in Indonesia from 2015-2025. The thematic maps compare the structural positions of co-word clusters based on standardised centrality and density scores. Centrality represents the extent to which a theme is connected to other themes in the research field, whereas density reflects the internal development and coherence of the theme. Each bubble represents a thematic cluster, with bubble size indicating the relative frequency of keywords associated with that cluster. Colours distinguish the clusters, while the abbreviated terms shown in the legend indicate the most representative keywords within each theme. The vertical and horizontal dashed lines represent the period-specific reference values used to divide the map into four quadrants: motor themes with high centrality and high density, niche themes with low centrality and high density, emerging or declining themes with low centrality and low density, and basic or transversal themes with high centrality but low density. The left panel presents themes identified during 2015–2019, whereas the right panel presents themes during 2020–2025, allowing changes in thematic importance, development, and connectivity to be examined over time.

**Table 2.**
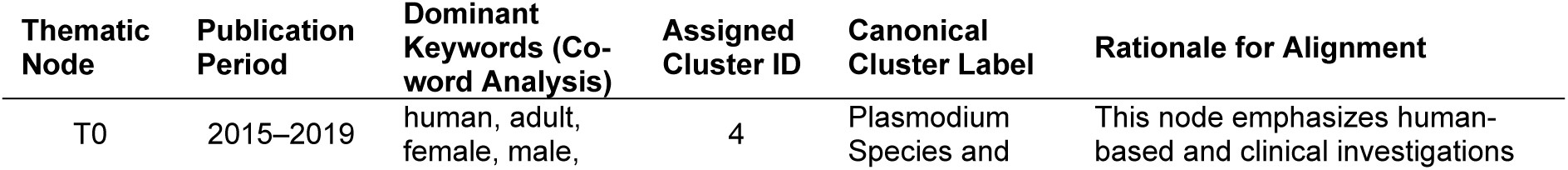
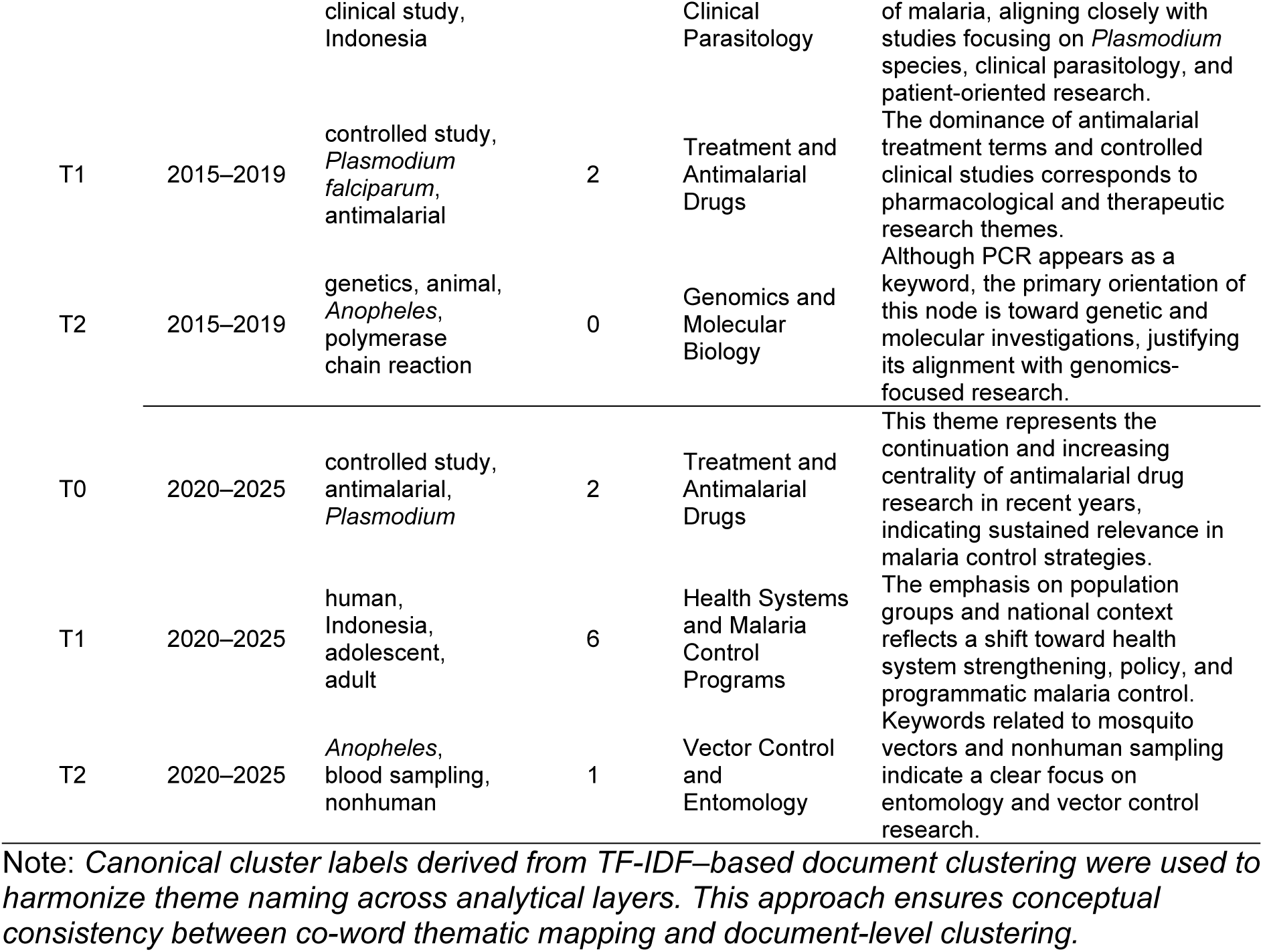
Alignment of co-word thematic map nodes with cluster-based research themes in Indonesian malaria research (2015–2025)

### Strategic thematic mapping

In the more recent period, the human-centred theme remained structurally prominent, with high centrality and density. However, its assignment to Health Systems and Malaria Control Programs should be interpreted cautiously because the dominant keywords primarily represent population and clinical characteristics rather than explicit programmatic or policy-related concepts. The treatment and antimalarial theme shifted toward a basic or transversal position, indicating that it remained broadly connected to other research areas but exhibited comparatively lower internal development. The Anopheles and non-human sampling theme remained in the emerging-or-declining quadrant, suggesting limited connectivity and thematic development within the overall research structure.

The findings indicate a reorganisation rather than a complete replacement of research priorities. Human-centred and clinical research remained prominent, while treatment-related research continued to serve as a broadly connected foundational theme.

Vector, non-human, and molecular-oriented topics remained comparatively peripheral. These results suggest increasing thematic differentiation within Indonesian malaria research, but they do not, by themselves, demonstrate field maturity, declining research activity, or future thematic prominence.

Fig 10 shows the relationship between cumulative citation impact and mean citations per document across themes in the corrected 586-publication thematic corpus. Treatment and Antimalarial Drugs recorded the highest total citation count (1,563) and the highest mean citation rate (22.65 citations per document). Plasmodium Species and Clinical Parasitology ranked second for both total citations (1,400) and mean citations per document (16.47), reflecting sustained scholarly influence.

**Fig 10.**
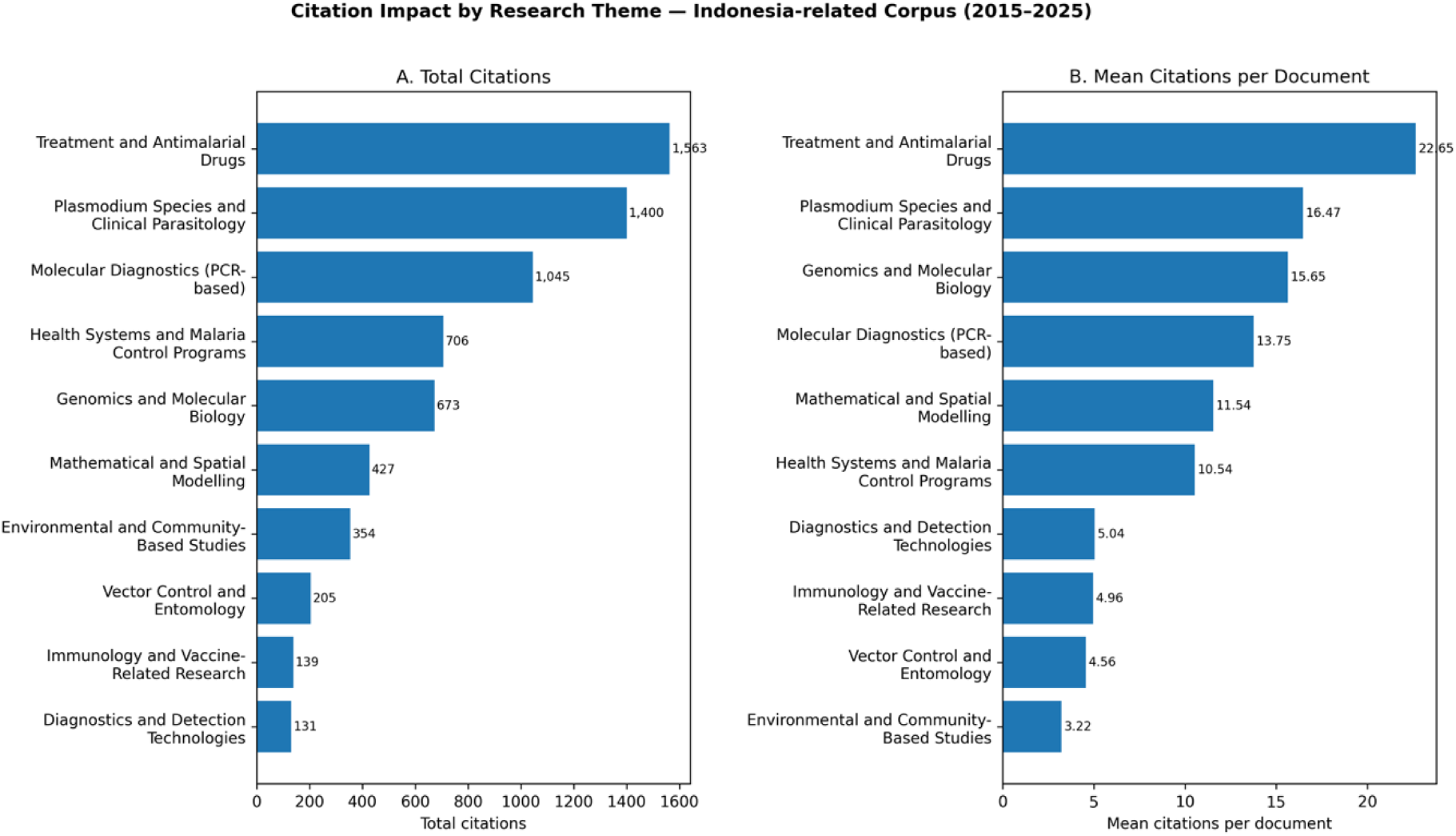
Citation analysis by theme in the final 586-publication thematic corpus. Treatment and Antimalarial Drugs recorded 1,563 total citations and 22.65 citations per document; Plasmodium Species and Clinical Parasitology recorded 1,400 and 16.47; Molecular Diagnostics (PCR-based) recorded 1,045 and 13.75; and Genomics and Molecular Biology recorded 673 and 15.65.

### Citation patterns by theme

Molecular Diagnostics (PCR-based) ranked third in total citations with 1,045 citations and a mean of 13.75 citations per document, whereas Genomics and Molecular Biology ranked third in mean citation intensity with 15.65 citations per document and accumulated 673 citations. Environmental and Community-Based Studies recorded 354 total citations and a mean of 3.22 citations per document, indicating that its growing publication prominence had not yet translated into comparable citation impact. Citation patterns therefore reflected different combinations of publication scale and per-document influence rather than a simple division between mature high-volume themes and specialised high-impact themes.

### Forecast of malaria research related to Indonesia (2026–2030)

Plasmodium Species and Clinical Parasitology are projected to increase gradually from approximately 9.5 publications in 2026 to 10.4 in 2030, while Molecular Diagnostics (PCR-based) increase from approximately 9.6 to 11.4. Treatment and Antimalarial Drugs remain nearly stable at approximately 5.9 publications annually, and Health Systems and Malaria Control Programs show only a slight increase from approximately 7.1 to 7.2 publications.

These forecasts are extrapolations of historical annual counts and should be interpreted as indicative trajectories rather than precise predictions. The models were based on only eleven annual observations and did not include prediction intervals; future funding, policy, epidemiological change, or technological developments may therefore produce different publication patterns.

Fig 11 presents exploratory forecasts for five leading themes using damped Holt linear-trend models fitted to annual publication counts from 2015 to 2025. Environmental and Community-Based Studies are projected to remain the most prominent of the modelled themes, increasing from approximately 17.5 publications in 2026 to 19.9 in 2030. This trajectory suggests continued research attention to environmental, behavioural, and community-level dimensions, although the forecast does not imply equivalent growth in citation impact.

**Fig 11.**
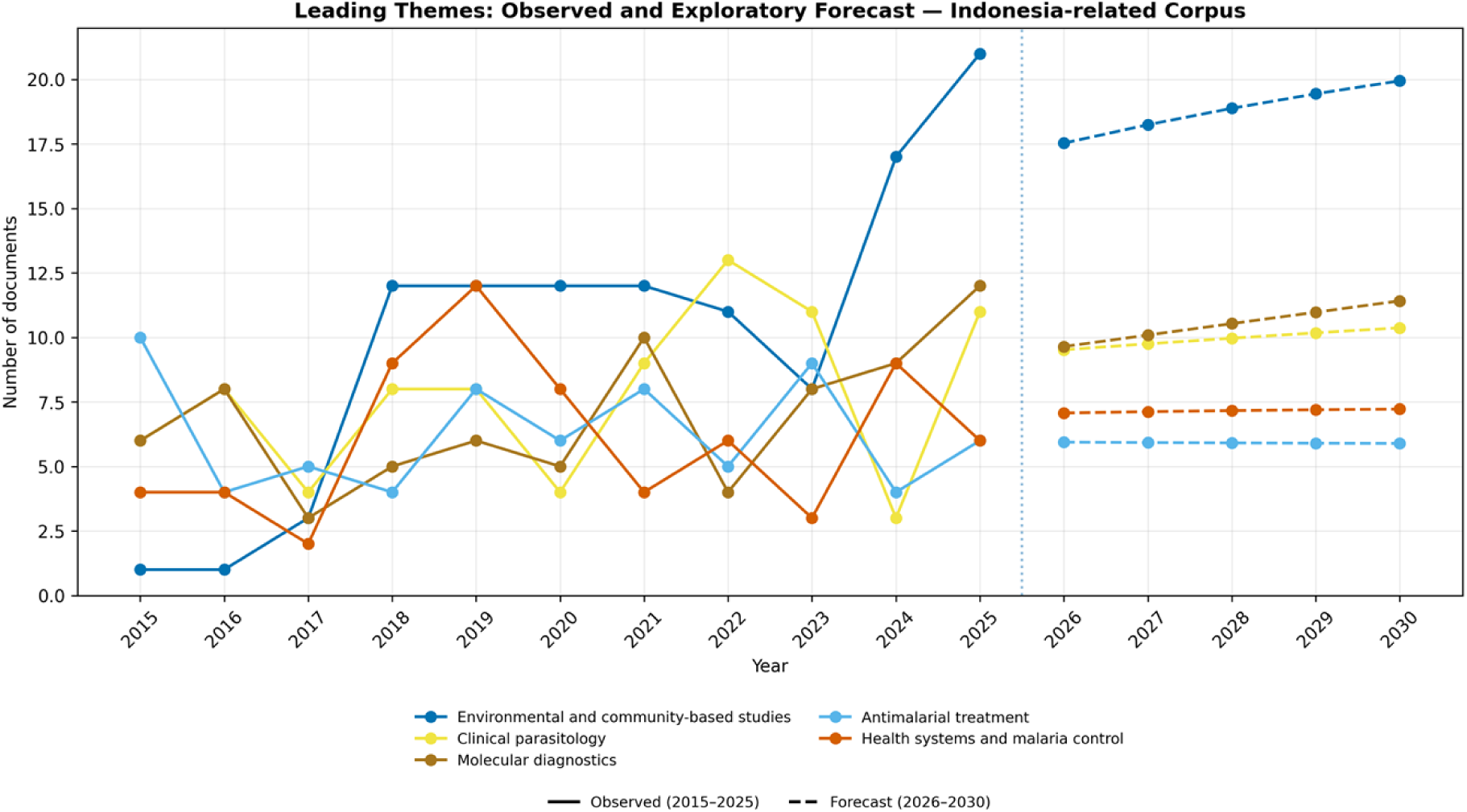
Exploratory forecast of leading malaria research themes related to Indonesia, 2026–2030. Damped Holt linear-trend models were fitted separately to annual publication counts from 2015 to 2025. Forecasts are indicative extrapolations and should not be interpreted as precise future values.

## Discussion

### Overall publication trend and field maturity

Among Southeast Asian countries, Thailand produced the largest number of Scopus-indexed malaria publications during 2015–2025, with 1,153 documents. Indonesia ranked second with 583 publications, followed by Malaysia with 466. Indonesia therefore produced fewer publications than Thailand but more than the other Southeast Asian countries included in the analysis. These differences should not be interpreted as direct evidence of national research quality or institutional performance, because publication output may also be influenced by research funding, international collaboration, disease burden, database coverage, and indexing practices.

Malaria research publication output in Indonesia showed an overall increase between 2015 and 2025, although the pattern was characterised by substantial annual fluctuations rather than uninterrupted growth. Publication output declined in 2017, increased markedly in 2018 and 2021, and reached its highest annual level in 2025. The cumulative publication curve showed continuous knowledge accumulation but did not display a clearly defined sigmoid pattern or plateau. Therefore, the observed trend does not provide sufficient evidence that Indonesian malaria research has reached a fully mature or stabilised phase. It more cautiously indicates sustained growth accompanied by increasing thematic diversification.

The changing thematic distribution may be interpreted in relation to Indonesia’s transition from controlling a relatively high malaria burden toward pursuing malaria elimination [34]. As transmission becomes increasingly concentrated in particular geographical areas, research priorities may increasingly address residual transmission, low-density infections, surveillance, and operational challenges [8,35]. National elimination strategies may also encourage greater attention to health systems, vector control, programme implementation, and surveillance-related research [8,9]. However, the bibliometric findings demonstrate changes in thematic prominence and should not be interpreted as direct evidence that national policy or funding changes caused the observed publication patterns.

Treatment and Antimalarial Drugs (Cluster 2) remained one of the most prominent research themes during the study period. Its continued presence indicates sustained scientific interest in antimalarial treatment, therapeutic effectiveness, resistance monitoring, and treatment optimisation [36]. This continuing relevance is consistent with the unresolved challenges associated with antimalarial drug resistance and variation in parasite and treatment-response profiles [37]. Nevertheless, its later position as a basic or transversal theme is more appropriately interpreted as evidence of broad connectivity with other research areas than as proof of thematic saturation or exclusively incremental development. Plasmodium Species and Clinical Parasitology (Cluster 4) also remained an important component of Indonesian malaria research. Its prominence reflects continued attention to parasite identification, clinical presentation, species differentiation, and diagnostic practice. The standardisation and harmonisation of diagnostic methods may facilitate the integration of clinical parasitology into epidemiological surveillance and applied disease-control research. Previous studies have emphasised the importance of coordinated diagnostic and surveillance approaches incorporating both human and animal health perspectives [38], consensus and standardisation in molecular diagnostic practice [39], integration of epidemiological sampling, diagnostic tools, and geospatial methods [40], and molecular approaches for parasite species characterisation [41]. However, these external studies provide contextual support and do not directly demonstrate that Indonesian clinical parasitology research has become fully mature or decentralised.

Environmental and Community-Based Studies (Cluster 5) became increasingly prominent in the temporal analysis, indicating growing research attention to environmental determinants, behavioural factors, community participation, and local transmission dynamics. These issues are relevant because malaria risk and intervention effectiveness are influenced by ecological, behavioural, and community-level conditions [42]. Health Systems and Malaria Control Programs (Cluster 6) also represented an identifiable theme within the ten-cluster analysis. External evidence supports the importance of programme implementation, health-service delivery, cross-sector collaboration, surveillance, policy development, and resource allocation in malaria control [42–45]. However, the human-centred node shown in the 2020–2025 strategic map primarily contains demographic and clinical keywords. Its position should therefore not be treated as direct evidence that health-system or policy research moved from the periphery to the centre of the thematic structure. Genomics and Molecular Biology (Cluster 0) and Immunology and Vaccine-Related Research (Cluster 7) remained identifiable but comparatively smaller thematic areas. In the strategic map, molecular, animal, Anopheles, and non-human research appeared relatively peripheral rather than clearly established as internally cohesive niche themes. These areas nevertheless remain scientifically relevant because molecular and immunological studies contribute to mechanistic understanding, parasite characterisation, therapeutic development, and longer-term innovation [46]. Their comparatively limited publication volume may also reflect the technological complexity, infrastructure requirements, cost, and extended development timelines commonly associated with molecular and vaccine-related research [47].

Vector-related research warrants continued attention because residual malaria transmission is influenced by ecological, behavioural, and vector-specific conditions that cannot be addressed through clinical treatment alone [48,49]. The increasing recognition of zoonotic malaria, including Plasmodium knowlesi in Southeast Asia and Indonesia, also reinforces the relevance of integrated human, animal, and vector surveillance [50].

Nevertheless, the strategic thematic map does not show a clear increase in the centrality of the vector-related node. It is therefore more accurate to describe vector research as a continuing and policy-relevant area rather than as a theme that has already moved to the centre of the Indonesian malaria research landscape.

The thematic findings indicate reorganisation and diversification rather than a complete replacement of previous research priorities. Clinical, parasitological, treatment-related, and molecular studies remained important, while environmental, community-based, and selected applied themes became more visible. The results do not demonstrate that biomedical or molecular research has become less important; instead, these areas continue to provide knowledge that may support surveillance, field diagnosis, treatment decisions, and integrated malaria-control strategies.

Artificial intelligence has increasingly been applied to disease detection and diagnostic-support systems [51]. Although numerous studies have explored AI-assisted malaria detection, the translation of these approaches into routine field implementation remains limited [52]. Within the Indonesian malaria corpus analysed in this study, AI-related research represented approximately 4% of publications and did not form a separate thematic cluster. This indicates that AI remains an emerging and embedded methodological approach rather than a dominant research theme in Indonesia [53].

### Intellectual influence and citation patterns

The citation analysis distinguishes cumulative scientific influence from citation intensity at the individual-document level. Treatment and Antimalarial Drugs recorded the highest total citation count and the highest mean number of citations per document, indicating that this theme combined substantial cumulative influence with strong citation performance at the publication level. Plasmodium Species and Clinical Parasitology ranked second for both total citations and mean citations per document, reflecting its sustained scholarly relevance within Indonesian malaria research.

Molecular Diagnostics (PCR-based) ranked third in total citations, whereas Genomics and Molecular Biology ranked third in mean citations per document. This difference indicates that molecular diagnostics generated a larger cumulative citation impact, while genomics-related publications achieved a slightly higher average citation intensity. Molecular Diagnostics therefore cannot be characterised as a theme with low cumulative influence, because it accumulated more than one thousand citations and maintained a comparatively high mean citation rate.

In contrast, Environmental and Community-Based Studies recorded a lower total citation count and a relatively low mean number of citations per document. Its increasing prominence in the temporal and forecasting analyses had therefore not yet translated into equivalent citation impact during the study period. This pattern may partly reflect the more recent expansion of the theme, although publication age, journal visibility, collaboration patterns, and disciplinary citation practices may also influence its citation profile.

Citation impact did not follow a simple distinction between high-volume mature themes and low-volume specialised themes. Treatment and clinical parasitology combined high cumulative and per-document citation impact, molecular diagnostics showed substantial cumulative influence, and genomics-related research demonstrated comparatively strong citation intensity despite a lower total citation volume. These complementary patterns may contribute differently to the scientific evidence required for malaria elimination, but citation performance alone cannot establish policy relevance, methodological quality, or thematic maturity [8].

### Research gaps and future priorities to support malaria elimination

The forecasting analysis provides an exploratory indication of publication trajectories for five prominent research themes between 2026 and 2030. Environmental and Community-Based Studies are projected to remain the most prominent theme and to show the largest continued increase in publication output. Plasmodium Species and Clinical Parasitology and Molecular Diagnostics (PCR-based) are also projected to increase gradually, indicating continued research attention rather than rapid expansion.

In contrast, Treatment and Antimalarial Drugs are projected to remain broadly stable throughout the forecast period. Health Systems and Malaria Control Programs also show only a slight increase and should therefore be interpreted as relatively stable rather than rapidly expanding. Vector Control and Entomology and surveillance-related research were not modelled as separate forecast series in Fig 10; consequently, their future growth cannot be inferred directly from the forecasting results. Nevertheless, these areas remain relevant to elimination-oriented research because low and spatially heterogeneous transmission requires effective programme implementation, vector management, and locally adapted interventions [49].

These projections are based on the continuation of historical publication patterns and should be interpreted as indicative trajectories rather than precise estimates of future output. The forecasting model does not establish the epidemiological, policy, technological, or funding mechanisms responsible for future changes. Developments in malaria transmission, national priorities, diagnostic technologies, or research funding could alter future trajectories beyond those represented by the model.

Artificial intelligence and machine learning represented approximately 4% of the Indonesian malaria research corpus and did not form a separate thematic cluster. Their application in diagnostic imaging, predictive modelling, and monitoring indicates emerging methodological relevance [54]. However, AI-related approaches currently function primarily as embedded analytical tools rather than as a major independent research domain. Their future contribution will depend on data availability, external validation, field implementation, interoperability with existing services, and evidence that these tools improve operational or clinical outcomes.

The forecasting results therefore suggest that future publication growth may be led primarily by Environmental and Community-Based Studies, with more gradual increases in Molecular Diagnostics and Clinical Parasitology. Recommendations concerning surveillance, vector control, health systems, or AI-assisted diagnosis should be presented as strategic research priorities informed by the wider thematic and epidemiological context, rather than as direct outputs of the forecasting model.

### Conceptual framework

We propose an interpretive framework linking three dimensions of Indonesian malaria research: thematic structure, scientific influence, and projected publication trajectory. The framework synthesises findings from temporal publication analysis, keyword clustering, strategic thematic mapping, citation analysis, and forecasting. It should be regarded as an analytical interpretation of the bibliometric results rather than as a validated causal model of research development.

The first dimension concerns thematic structure. Human-centred clinical research remained a structurally prominent motor theme across both study periods. Treatment and Antimalarial Drugs also remained an important foundational theme, although its later position was more consistent with a basic or transversal theme than with a highly developed motor theme. Environmental and Community-Based Studies showed increasing proportional and projected publication prominence, but their comparatively low citation performance indicates that increasing research activity had not yet resulted in equivalent scholarly influence.

The second dimension concerns scientific impact. Treatment and Antimalarial Drugs and Plasmodium Species and Clinical Parasitology combined high cumulative citation counts with high mean citations per document. Molecular Diagnostics generated substantial cumulative citation impact, whereas Genomics and Molecular Biology demonstrated comparatively high citation intensity despite a lower overall citation volume. Scientific influence therefore arose from different combinations of publication scale and average citation performance.

The third dimension concerns projected publication trajectories. Environmental and Community-Based Studies are expected to show the strongest continued growth, while Molecular Diagnostics and Plasmodium Species and Clinical Parasitology are projected to increase more gradually. Treatment and Antimalarial Drugs and Health Systems and Malaria Control Programs are projected to remain relatively stable. These forecasts identify possible directions of future publication activity but do not demonstrate thematic saturation, causal policy effects, or future research quality.

Taken together, the framework indicates that Indonesian malaria research contains established clinical and treatment-related foundations, expanding environmental and community-oriented research, and selected molecular themes with substantial citation influence. Future research prioritisation should therefore consider thematic relevance, scientific impact, epidemiological need, implementation feasibility, and potential contribution to malaria elimination rather than publication volume alone.

These findings are relevant to several groups involved in malaria research and elimination in Indonesia. Researchers may use the identified thematic patterns and evidence gaps to guide future study design and collaboration, while research funders may use them to identify areas that are well established or comparatively underdeveloped. National and subnational malaria programme managers and policymakers may find the mapping of environmental, community, diagnostic, treatment, and health-system research useful when aligning research priorities with elimination needs. The results may also inform laboratory and public-health practitioners by highlighting the continued importance of diagnostic implementation, surveillance, treatment monitoring, and locally adapted interventions.

However, the bibliometric findings describe patterns in the published evidence base and should not be interpreted as direct evidence of intervention effectiveness or policy impact.

### Limitations

This study has several limitations. First, the analysis was restricted to Scopus-indexed publications and may therefore omit relevant research published in non-indexed Indonesian journals or grey literature. This could underrepresent locally focused community, programme, or policy research.

Second, the review used related but distinct analytical denominators. The initial 592-record Indonesia-candidate set underwent a final relevance audit that excluded four false-positive records, yielding 588 eligible Indonesia-related publications. Affiliation-based country comparisons used 583 records with an Indonesian affiliation identifiable in the exported Scopus Affiliations field, whereas keyword-based thematic analyses used 586 eligible records because two publications lacked usable author and index keywords. These definitions were applied for different analytical purposes, but absolute counts should not be compared across components without considering the relevant denominator.

Third, bibliometric analyses depend on the completeness and consistency of titles, abstracts, keywords, affiliations, and citation metadata. Thematic clustering based on TF–IDF and K-means captures textual similarity rather than the complete substantive content of each publication, and interdisciplinary studies may overlap across themes. The low silhouette values and disagreement among internal validation metrics also indicate that the ten-cluster solution should be interpreted as a pragmatic thematic representation rather than a uniquely optimal structure.

Fourth, citation counts were used as proxies for intellectual influence but do not directly measure methodological quality, societal benefit, or policy impact. Citation patterns are also affected by publication age, journal visibility, collaboration, and disciplinary citation practices.

Finally, the strategic thematic maps involve interpretive alignment between period-specific co-word nodes and the separate ten-cluster document model. Nodes from 2015–2019 are not necessarily direct equivalents of those from 2020–2025. Forecasts were based on damped Holt trend extrapolation from only eleven annual observations and did not include prediction intervals. They should therefore be treated as exploratory indications rather than precise estimates of future output.

## Conclusion

Malaria research related to Indonesia showed overall growth and increasing thematic diversification between 2015 and 2025. Clinical parasitology and antimalarial treatment remained important foundations, while environmental, community-based, diagnostic, and implementation-oriented studies became increasingly prominent. The findings indicate a reorganisation of priorities rather than a complete shift away from biomedical research. A balanced agenda that integrates established clinical and laboratory approaches with applied, community-level, and health-system research may better support Indonesia’s malaria elimination efforts.

## Author contributions

Budi Setiawan (BS): Conceptualization, Methodology, Formal analysis, Writing – original draft, Visualization.

Jonathan M. Cooper (JMC): Supervision, Writing – review & editing. Julien Reboud (JR): Supervision, Writing – review & editing.

## Data availability

The Scopus records used in this study are third-party bibliographic data available through Scopus under its standard access conditions. Derived datasets underlying the reported analyses are provided as S1 and S2 Data, and the complete analysis script is provided as S1 Code. Abstract text licensed by Scopus is not redistributed.

## Supporting information

**S1 Data.** Eligible Indonesia-related publication dataset used for descriptive analyses (n = 588).

**S2 Data.** Keyword-based thematic analysis dataset with final cluster assignments (n = 586).

**S1 Code.** Python script used to finalise the corpus and reproduce the corrected analyses and figures.

